# NN50 and pNN50, two time-domain heart rate variability parameters were associated with 30-day all-cause mortality in patients admitted to intensive care unit: A Retrospective Study of the MIMIC-IV Database

**DOI:** 10.1101/2023.03.09.23287074

**Authors:** Sheran Li, Qiyu Yang, Peiyu Wu, Yujing Lu, Zhengfei Yang, Longyuan Jiang

## Abstract

**Objective:** Our study aims to evaluate the association between heart rate variability (HRV) and short and long-term prognosis in patients admitted to intensive care unit (ICU).

**Methods and Results:** Adult patients continuously monitored for over 24h in ICUs from the MIMIC-IV Waveform Database were recruited in our study. Twenty HRV-related variables (8 time-domain, 6 frequency-domain; and 6 nonlinear variables) were calculated based on RR intervals. The association between HRV and 30-day all-cause mortality was assessed. Ninety-three patients met the inclusion criteria and were classified into 30-day survivor group and non-survivor groups based on their survival status. The 30-day all-cause mortality rate was 17.2%. NN50 and pNN50 were both significantly higher in non-survivors compared to survivors, whereas the rest of the time-domain, frequency domain and non-linear HRV parameters did not differ significantly between the two groups (all *P* >0.05). In addition, at 180 days after admission, non-survivors had significantly higher levels of NN50 and rMSSD than the survivors. However, NN50 was not an independent predictor of 30-day all-cause mortality in patients by multivariate COX regression analysis (HR, 1.0; 95% CI, 1.000 - 1.001; *P* =0.594). The Area Under the Curve (AUC), cut-off value, sensitivity and specificity of NN50 for predicting 30-day all-cause mortality using ROC were 0.67, 799, 0.813 and 0.584, respectively. Plotting Kaplan-Meier analysis using this cut-off value showed that patients with high NN50 had considerably greater 30-day all-cause mortality than those with low NN50 (*P* < 0.001).

**Conclusion:** NN50 and pNN50 are associated with elevated 30-day all-cause mortality in ICU patients but are not independent predictors of all-cause mortality using multivariate COX regression analysis.

## Introduction

Reported all-cause mortality in intensive care unit (ICU) patients ranges from 10% to 60%[1] and is associated with multiple variables including but not limited to the age, type and severity of the primary illness[2]. Heart rate variability (HRV), defined as variations between consecutive heartbeats or R-R intervals on electrocardiogram (ECG) recordings, reflects the balance between sympathetic (SNS), parasympathetic nervous system (PNS) and endocrine system activities in modulating the heart[3]. It has long been recognized as an important predictor of clinical conditions such as fetal distress[4] and myocardial infarction[5]. Changes in HRV have also been reported in ICU patients, reflecting the presence of cardiac autonomic dysfunction[3]. However, different studies have reported different HRV variables as predictors of disease severity or mortality in critical care patients. For example, Schmidt et al. found that a reduction in very low-frequency power was predictive of 28-day mortality in patients with multiple organ dysfunction syndrome (MODS)[6], whereas Pontet et al. reported that the low-frequency spectral component was the best predictor of the development of MODS[7]. In septic patients, Chen et al. found that SDNN (standard deviations of RR intervals) and normalized high-frequency power were valuable predictors of in-hospital mortality [8], while Samsudin et al. found that the combination of mean RR interval, detrended fluctuation analysis (DFAa2), and age, systolic blood pressure (SBP), and respiratory rate outperformed the traditional quick sequential organ failure assessment (qSOFA) score in predicting 30-day mortality, ICU admission and intubation [9]. The differences in these studies, which may be explained by sample size, duration of ECG recordings, etc., suggest that a clinically robust HRV indicator is still lacking. This knowledge gap may be filled with the future full release of the American Medical Information Mart for Intensive Care (MIMIC)-IV Waveform Database, which will include approximately 10,000 ECG records from ICU patients admitted to the Beth Israel Deaconess Medical Center (BIDMC) between 2008 and 2019, providing ECGs from a much larger cohort of ICU patients for HRV-related research. Our study is a preliminary analysis of the MIMIC-IV Waveform Database (version 2.2), which includes a total of 198 patients and aims to delineate the association between HRV variables and short- and long-term prognosis in ICU patients.

## Methods

### Study Participants

The patients in this study are a subset of patients from the MIMIC-IV (v2.2) database, a freely available database created and maintained by the MIT Computational Physiology Laboratory[10]. The database contains information of all patients being treated in either the emergency department or an ICU of Beth Israel Deaconess Medical Center (BIDMC) between the years 2008 and 2019. Every patient’s admission details, vital signs, diagnoses, laboratory values, microbiology measurements, medication treatment, date of death (if applicable), and other comprehensive information were recorded. Patients in our study had bedside cardiac monitoring data available, which is stored in a separate database called the MIMIC-IV Waveform Database [11] (https://physionet.org/content/mimic4wdb/0.1.0/). This database contains real-time monitoring and assessment of patients in ICUs using ECGs, photoplethysmograms, blood pressure monitors, and beyond. The two databases are linked by unique patient IDs. All personal information in the database is de-identified, so informed consent and ethics approval are not required to use the data for research purposes. These two databases are available on the official PhysioNet website(https://physionet.org/) to researchers who complete the Collaborative Institutional Training Initiative (CITI) course and pass both the “Conflicts of Interest” and “Data or Specimens Only Research” tests. Sheran Li, as the principal investigator, was granted the right to extract data (Pass ID:51880407). The research was also approved by the Ethics Committee of Sun Yat-sen Memorial Hospital, Sun Yat-sen University (ID: SYSKY-2023-089-01).

### Study Protocol

Patient characteristics collected included demographics (age, sex, race and marital status), vital signs [systolic (SBP) and diastolic blood pressure (DBP), mean arterial pressure (MAP), heart rate (HR), and oxygen saturation (SpO2)], comorbidities and laboratory values. The comorbidities analyzed included sepsis, infections mainly pulmonary and urinary tract infections, hypertension, diabetes (type 1 and type 2), heart failure including diastolic and systolic heart failure, renal failure, including acute and chronic renal failure, respiratory failure, hepatic failure, stroke and transient ischemic attack (TIA), malignancy, atrial fibrillation (AF), myocardial infarction, coronaryangioplasty implant and graft, lipid disorders including hypertriglyceridemia and hypercholesterolemia, obesity and malnutrition. Laboratory values analyzed included red (RBC) and white blood cell (WBC), platelets, hemoglobin, serum glucose, serum creatinine, blood urea nitrogen (BUN), anion gap, serum potassium, serum sodium, serum calcium, serum magnesium, serum chloride and serum bicarbonate. Data extraction was performed using iPython software (v8.6.0) through self-developed codes. The extracted data were reviewed and manually selected for final analysis. For example, in patients with multiple complete blood counts during the ICU stay, the result of the test before or during the monitoring period was used for the final analysis. A similar strategy was used for all other variables to represent the clinical picture as accurately as possible. Variables were excluded if they had more than 3% missing values, such as height, albumin, alanine aminotransferase, aspartate aminotransferase, total bilirubin, lipid profile, arterial blood gas results, lactate, absolute cell count and percentage of blood basophils, eosinophils, lymphocytes, monocytes and neutrophils Therefore, no selection bias was present.

### Evaluation of time-domain, frequency-domain; and nonlinear variables of HRV

Analysis was performed using Python and Matlab software provided by PhysioNet [12, 13]. . A detailed description and explanation of all HRV variables is provided in Supplementary Table 1. The original ECG signal was processed using double median filter and baseline shift removal, after which a biorthogonal 4.4 wavelet filter was applied to derive the QRS waves. A moving window integration filter was then used to remove body motion artifacts and to detect R peaks. RR intervals between 250 and 1600 ms were selected criteria for sinus rhythm. All time-domain variables were calculated accordingly. For the frequency-domain variables, a Burg algorithm was used to estimate the auto-regression model (AR) spectrum [14], and the spectral power of the ultra-low-frequency (ULF), very-low-frequency (VLF), low-frequency (LF), high-frequency (HF) range, the total power (TP, see Supplementary Table 1 for more details), and the ratio of LF to HF power were determined. All power measurements were expressed in ms^2^.

For nonlinear variables, the Poincaré plot was calculated and SD1 and SD2 were determined as previously described[15]. Approximate entropy (ApEn) and sample entropy (SampEn) were determined with a parameter of m = 2 and similarity criterion = 20% of SD as previously described[16, 17].. Detrended fluctuation analysis (DFA) was performed and DFA (α1) and DFA (α2) were determined using a previously established method[18].

All variables were calculated from the entire RR intervals and expressed as the average value for the entire 24-hour recording period.

Included patients were classified as survivors or non-survivors based on their survival status at 7, 30, 90, 180 and 365 days after admission. The main text includes all results at 30 days and partial results at 7 days and 180 days after admission. Comparison between survivors and non-survivors at the other time points are available in the Supplementary Appendix.

### Statistical Analysis

Data analysis was conducted using GraphPad Prism (v9.0.0) and SPSS (v26). Normality of the data was checked using the Kolmogorov-Smirnov test. Results were expressed as mean ± SD for normally distributed variables, median (first quartile-third quartile) for non-normally distributed continuous variables, and number (%) for categorical variables. Continuous variables were compared by t-test for normally distributed variables or by Mann-Whitney U test for non-normally distributed variables. Pearson’s chi-squared test and Fisher’s test were used to compare categorical variables. Variables that were significantly different between survivors and non-survivors were included in univariate Cox regression analysis. Multicollinearity was assessed, and variables that were correlated with each other were individually included in the multivariate Cox regression model. Multivariate Cox regression analysis was then used to identify independent predictors of mortality after adjustment for age, sex and/or other potential confounders identified by univariate Cox regression analysis. Receiver Operating Characteristic (ROC) analysis was used to assess the predictive ability of the identified variables for 30-day mortality, the sensitivity and specificity of each variable, and the Area Under Curve (AUC). The Youden index was applied to determine the best cut-off level of the identified variables, which was then utilized to classify the patients into high and low-level groups. The Kaplan-Meier (KM) survival curves for the two groups were plotted and compared using the log-rank test.

Significance was defined as a *P* value < 0.05 for all analyses.

## Results

### Patient Characteristics

Table 1 shows patients’ baseline characteristics and comparisons of variables between survivors and non-survivors at 30 days after admission. Of the 198 patients in the database, 22 patients had missing demographic information and 83 patients had ECG recordings of less than 24 hours, which were excluded from the final analysis (Supplementary Figure 1). No significant difference was found between included and excluded patients in our primary variables of interest (all-cause mortality,) except for ICU stay, presence of respiratory failure, body weight and calcium level (Supplementary Table 2), suggesting that the missing of the data were random. After exclusion, our study included 93 patients with an average age of 64.6 ± 15.8 years and a median ICU stay of 3.0 (1.9-9.2) days. In the study cohort, 45 patients (48.4%) were female, and the majority of the participants were Caucasian (61.3%), with only one Asian patient included. 34.4% of the patients were married. A total of 16 patients (17.2%) did not survive to 30 days after admission.

**Table 1.**
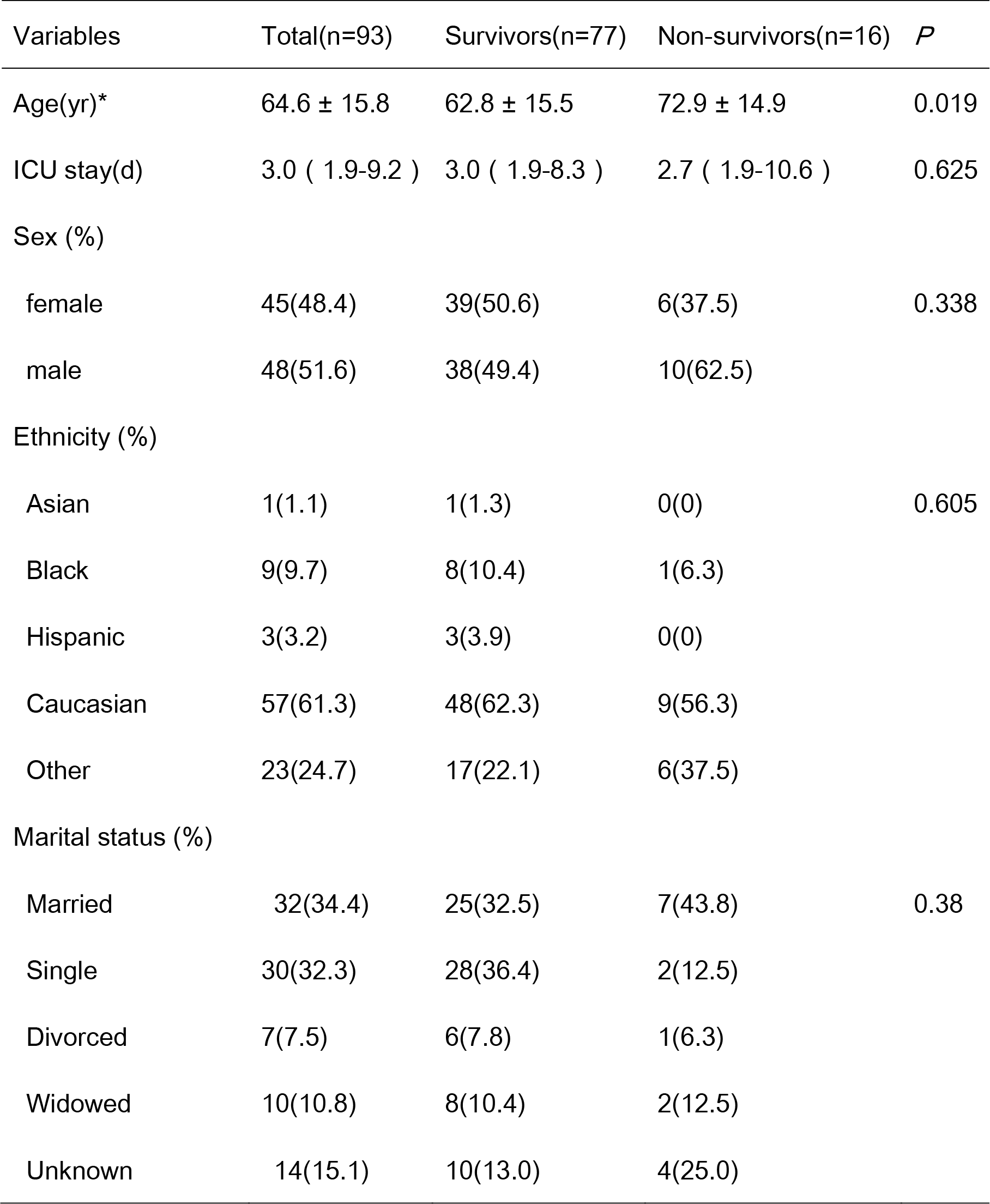
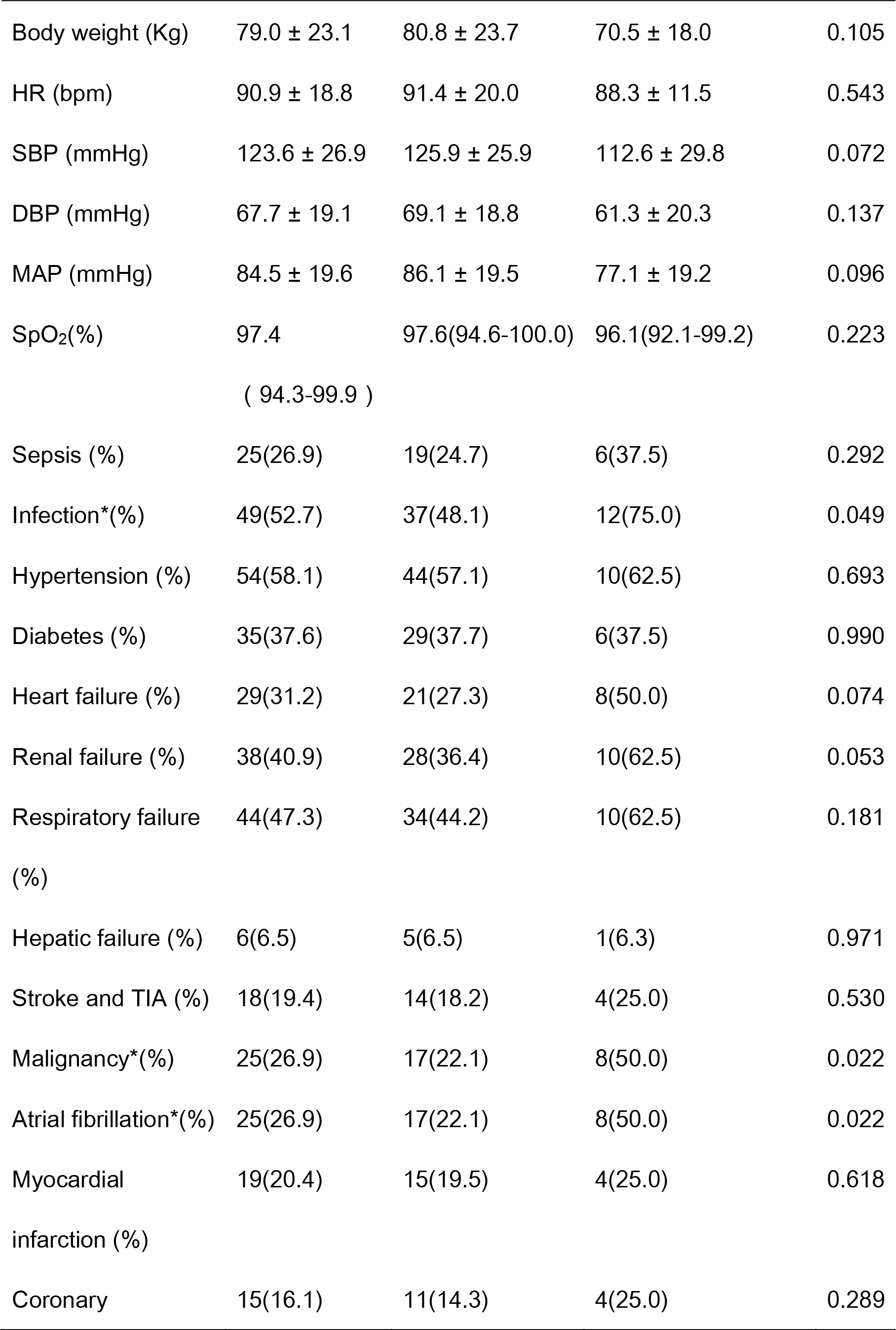
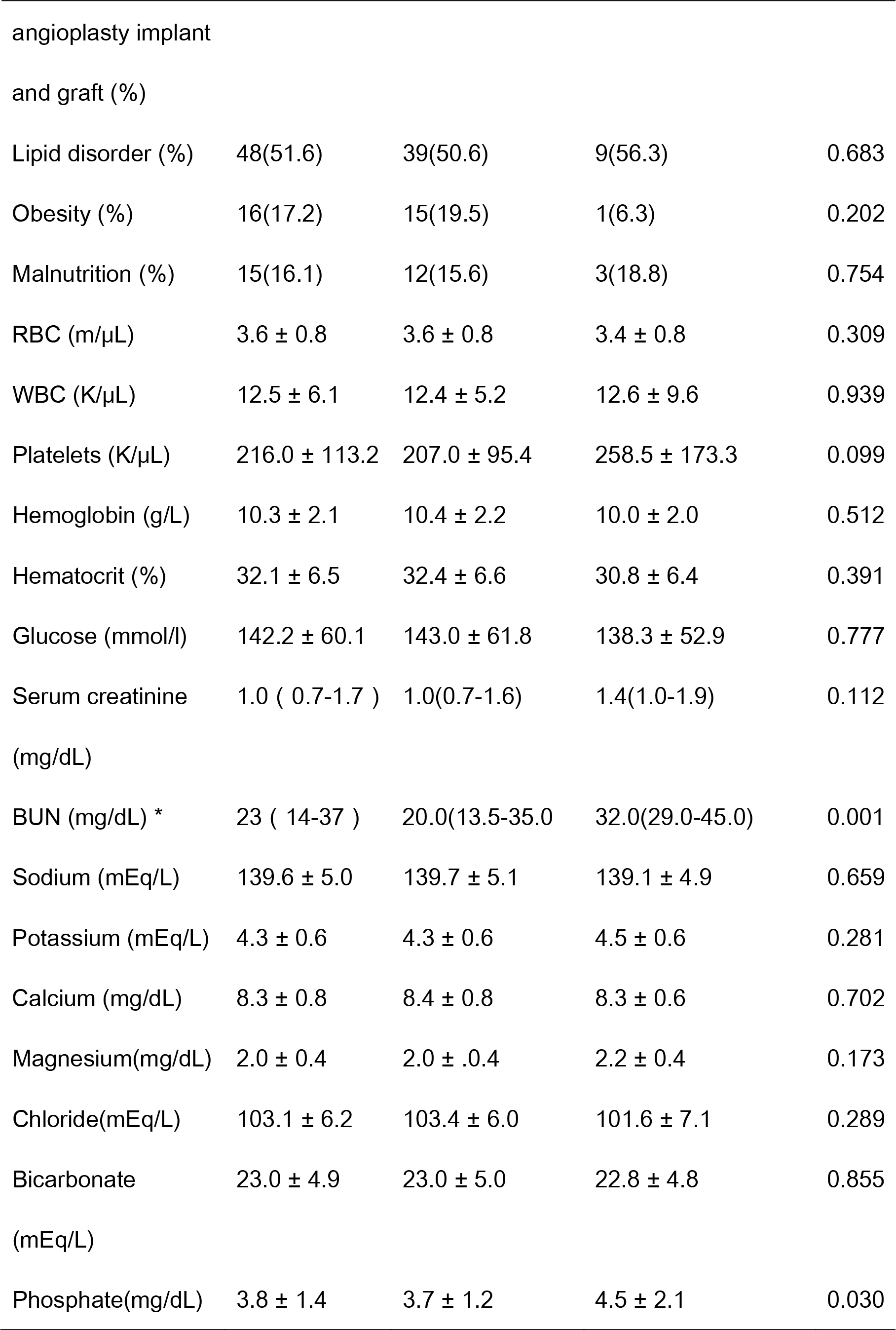
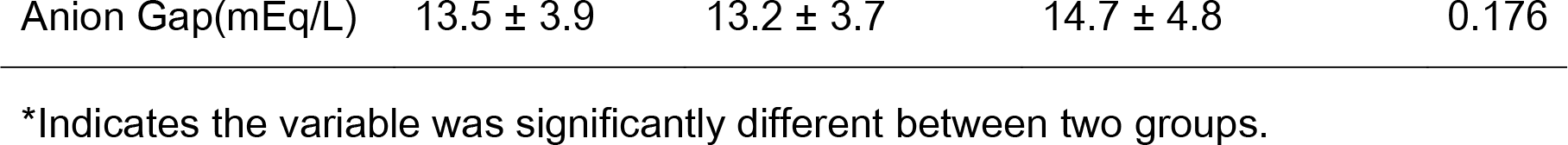
Patient characteristics in the survivor and non-survivor group at 30 days

Survivors were significantly younger than non-survivors (*P* <0.05). Vital signs including HR, SBP, DBP, MAP and SpO2 did not differ between the survivor and non-survivor groups (*P* > 0.05). The prevalence of hypertension, the most common comorbidity in the study cohort, did not differ between the two groups (*P* > 0.05). On the other hand, the prevalence of infection, present in 52.7% of the study population, was substantially greater in non-survivors than in survivors (75.0% vs. 48.1%, *P* = 0.049). The non-survivor group also had a significantly greater prevalence of malignancy, and atrial fibrillation (all *P* <0.05). The prevalence of sepsis, diabetes, heart failure, renal failure, respiratory failure, hepatic failure, stroke and TIA, myocardial infarction, history of coronaryangioplasty implant and graft, lipid disorder, obesity and malnutrition did not differ between survivors and non-survivors (all *P* > 0.05).

Laboratory tests showed that BUN and blood phosphate levels were significantly greater in non-survivors than in survivors [BUN,32.0(29.0-45.0) vs 20.0 (13.5-35.0) mg/dL, *P* = 0.001; blood phosphate,4.5 ± 2.1 vs. 3.7 ± 1.2 mg/dL, *P* = 0.030], while the remaining covariates including RBC, WBC, platelets, hemoglobin, hematocrit, serum glucose, creatinine, sodium, potassium, calcium, magnesium, chloride, bicarbonate and anion gap did not differ between survivors and non-survivors (*P* > 0.05).

Comparison of variables between the survivor and non-survivor groups at 7, 90, 180 and 365 days is shown in Supplementary Tables 3, 4, 5 and 6, respectively.

Supplementary Table 7 summarizes the variables that differed significantly between the survivor and non-survivor groups at these time points. BUN was the only variable that was significantly different between the two groups at all time points.

Older age was associated with increased 7-, 30-, 90- and 180-day mortality.

### Comparison of HRV variables between the survivor and non-survivor groups at 30 days after admission

Table 2 shows the comparison of HRV parameters between the two groups.

**Table 2.**
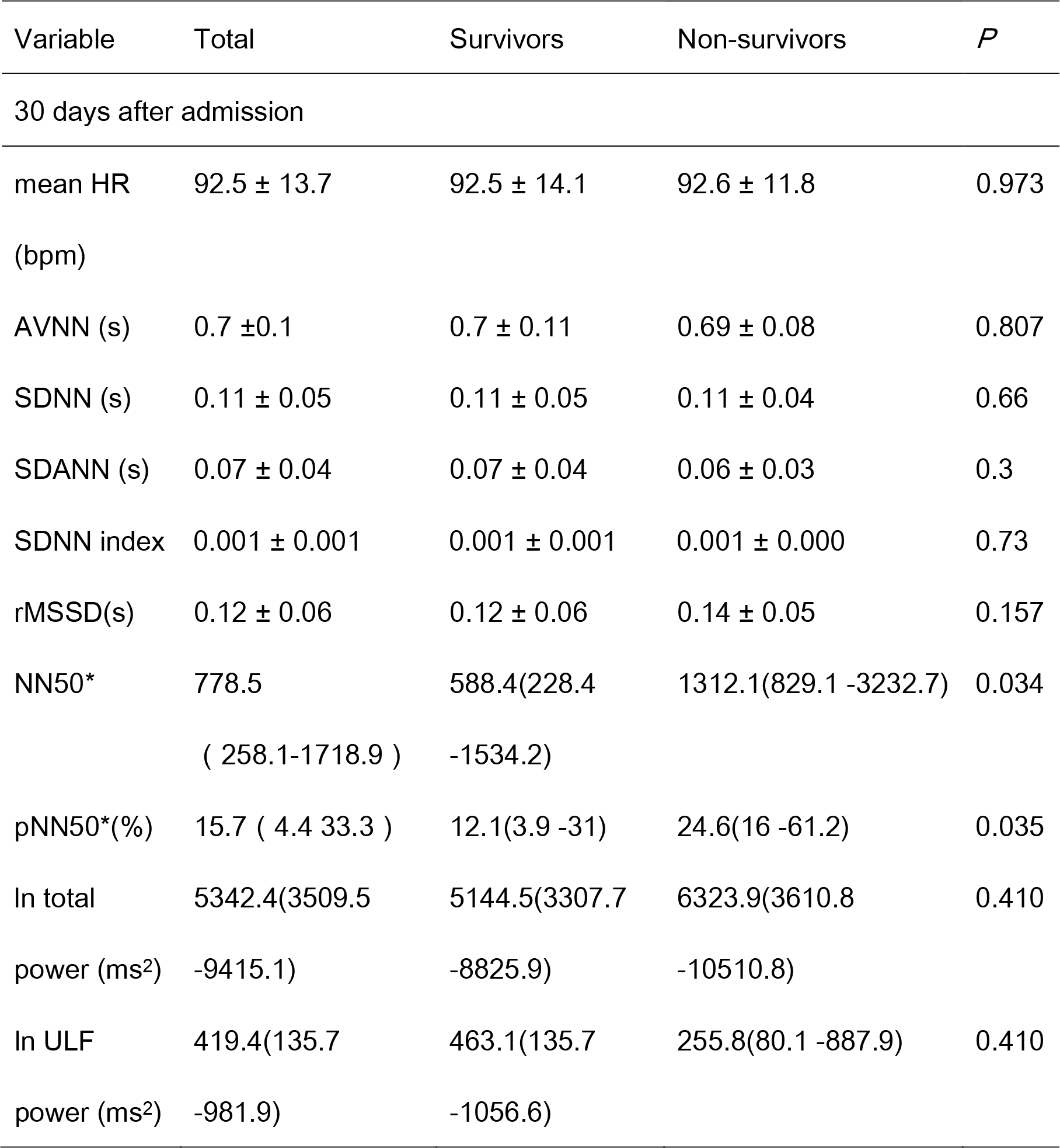
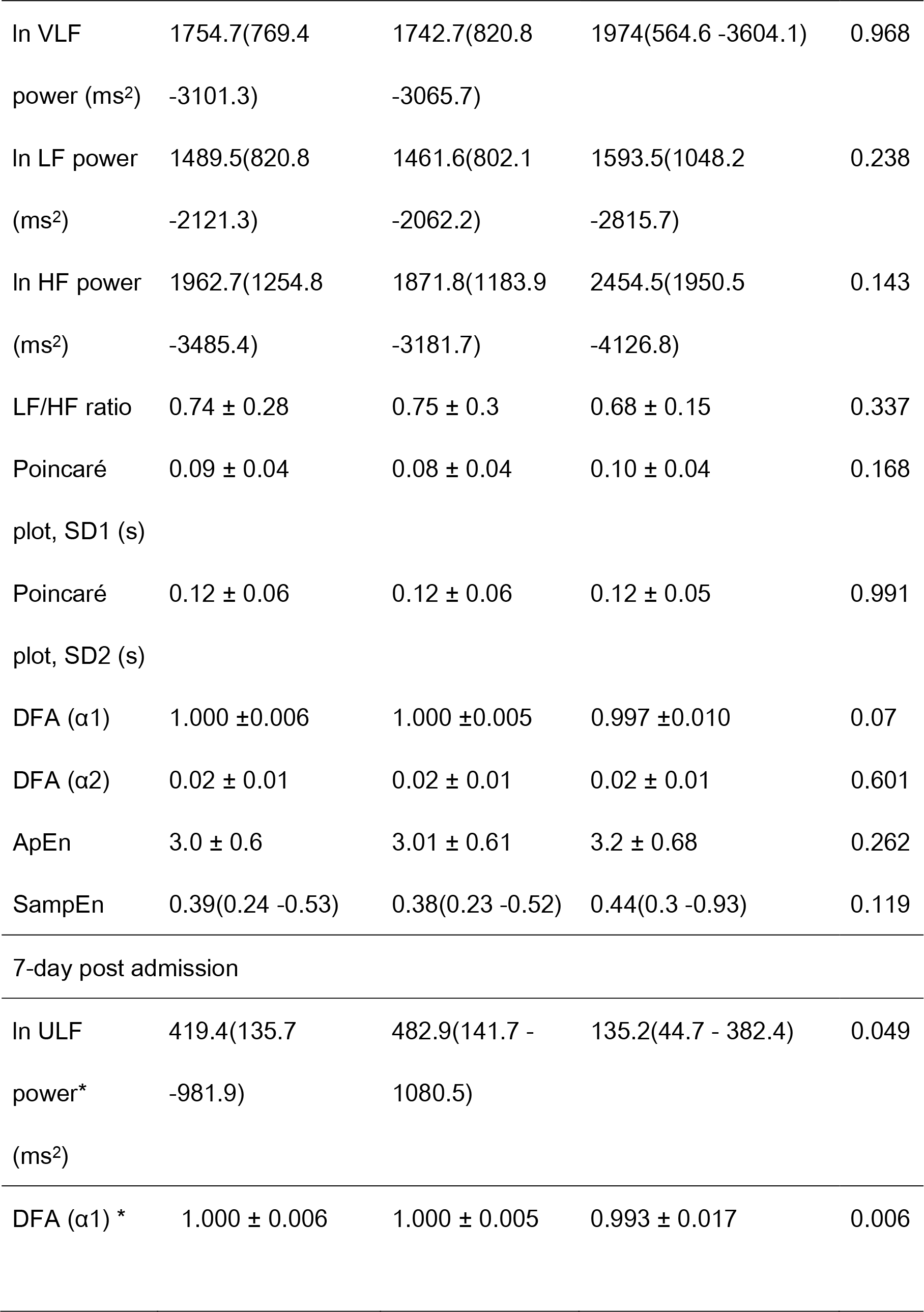
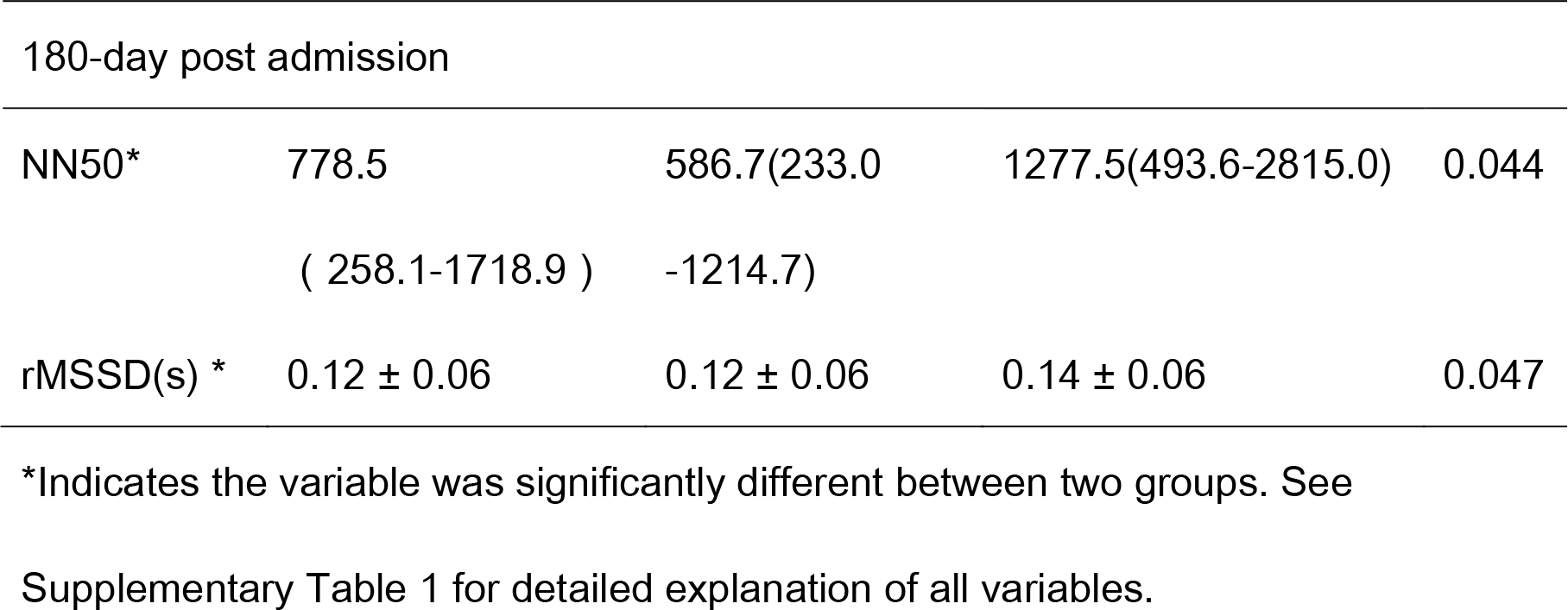
Comparisons of HRV variables between survivors and non-survivors at 30 days after admission

Non-survivors had a substantially greater level of NN50 and pNN50 than the survivors [NN50, 1312.1(829.1 -3232.7) vs.588.4(228.4 -1534.2), *P*=0.034; pNN50, 24.6(16 -61.2) vs.12.1(3.9 -31) %, *P*=0.035]. Otherwise, no significant difference was found between the remaining time-domain, frequency domain and nonlinear HRV parameters (all *P* >0.05).

In addition, non-survivors had a significantly lower level of ULF power and DFA (α1) at 7 days after admission compared to the survivors (all *P*<0.05), whereas they had a significantly higher level of NN50 [1277.5(493.6-2815.0) vs. 586.7(233.0 -1214.7), *P*=0.044] and rMSSD (0.14 ±0.06 vs.0.12 ±0.06s, *P*=0.047) at 180 days after admission.

### Univariate and Multivariate Cox Regression Analyses of HRV Variables and Potential Confounders for Prediction of 30-Day All-Cause Mortality

Table 3 shows the HR and 95% CI of variables that differed significantly between the survivor and non-survivor groups for predicting 30-day all-cause mortality by univariate Cox regression analysis. The result showed that unadjusted age, presence of malignancy and AF, BUN, phosphate, NN50 and pNN50 were all associated with increased 30-day all-cause mortality in ICU patients (*P*<0.05).

**Table 3.**
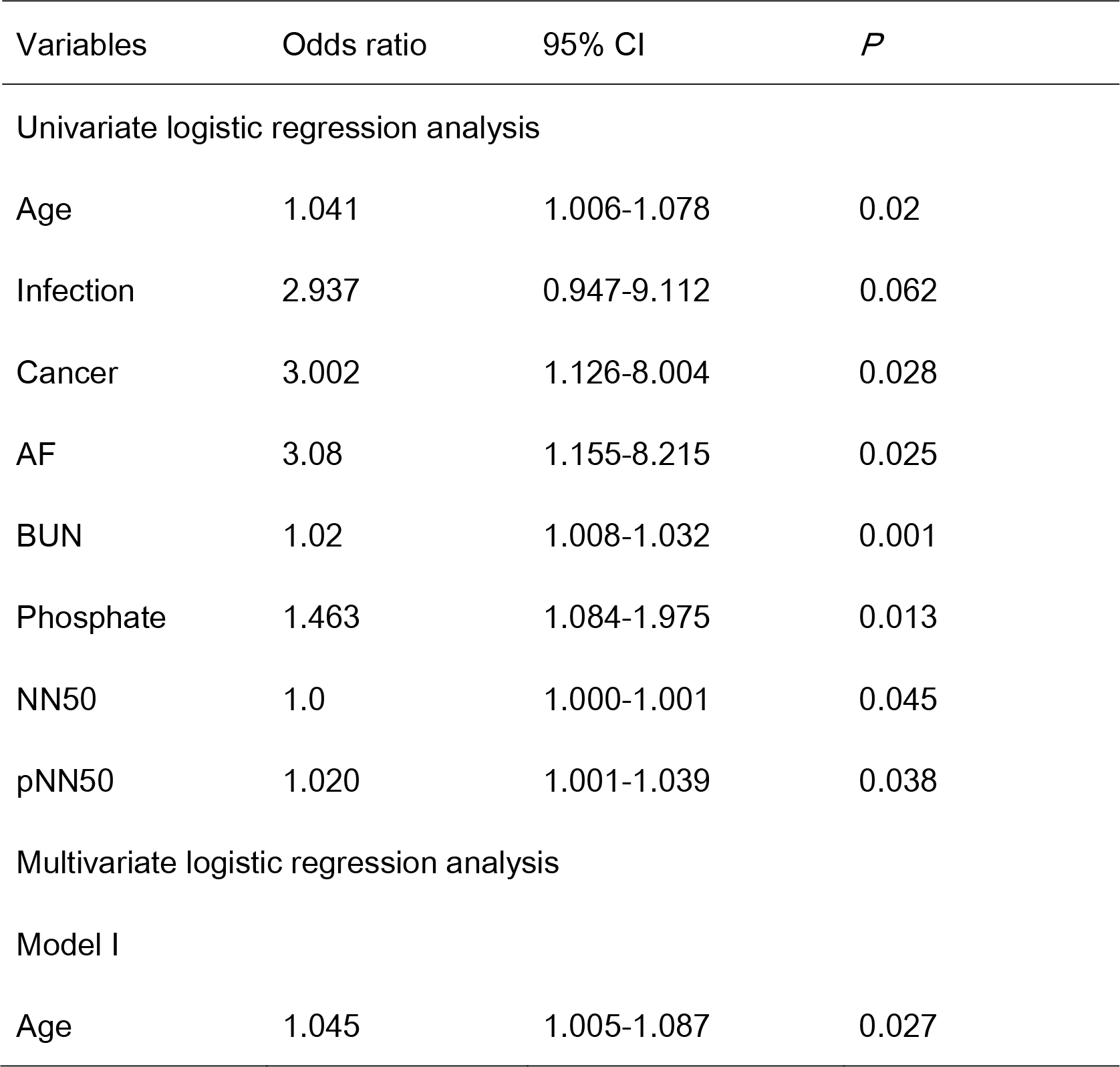
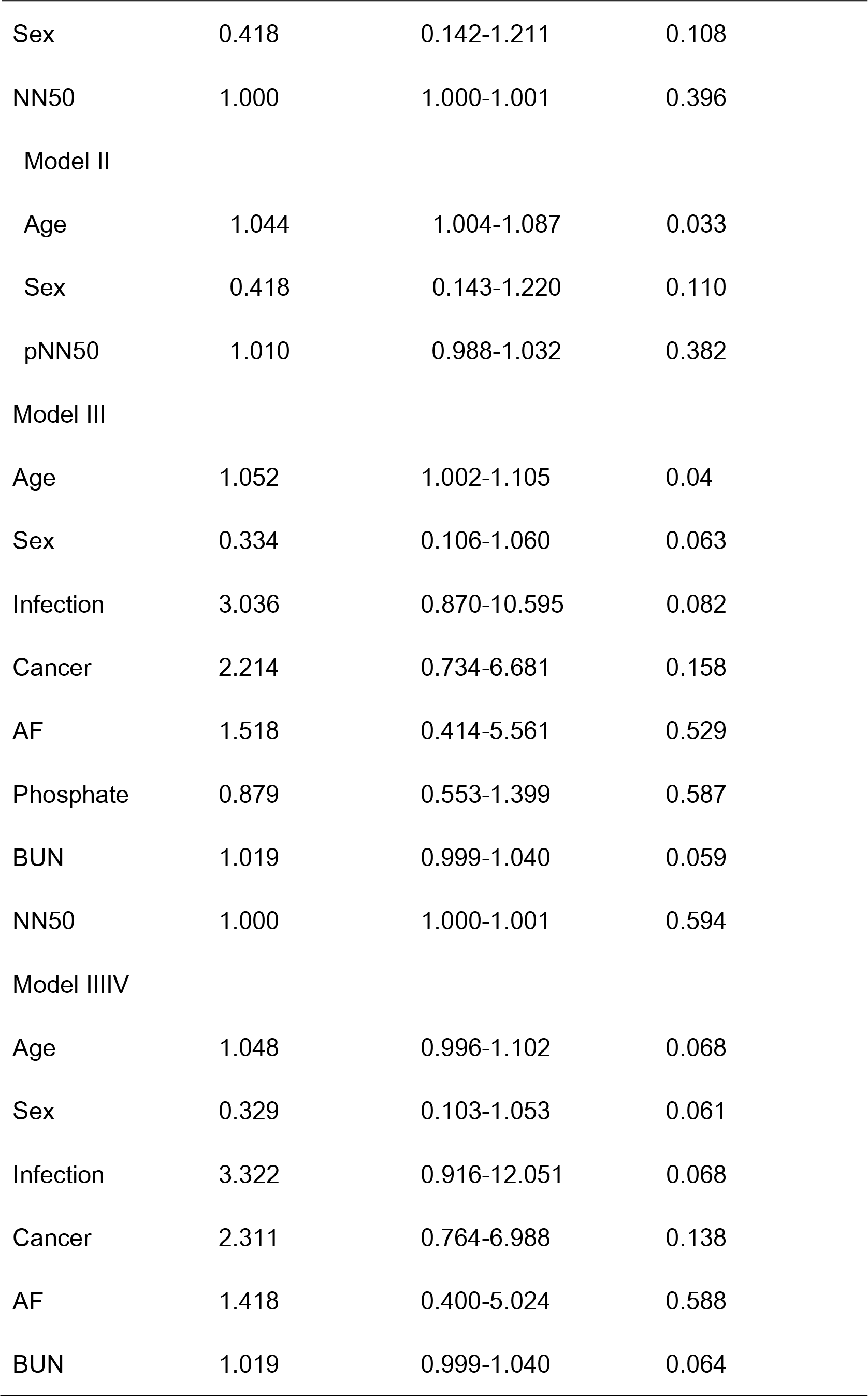
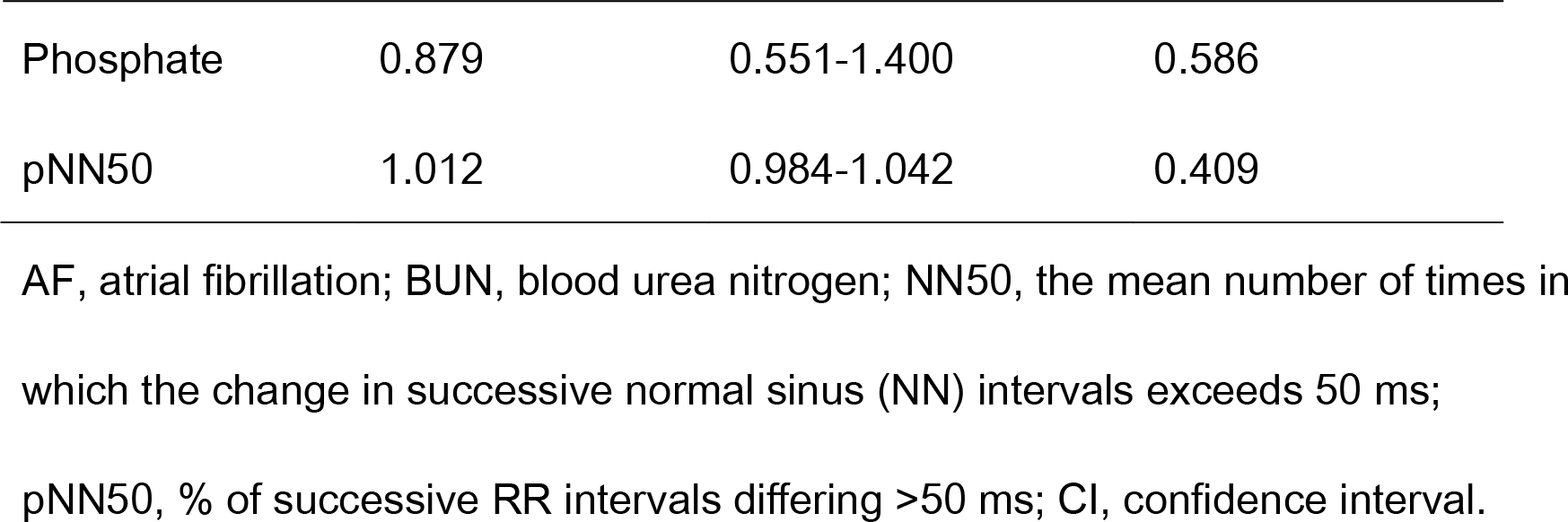
Univariate and Multivariate COX analysis of risk factors for 30-day all-cause mortality in patients by logistic regression analysis.

| Variables | Odds ratio | 95% CI | *P* |
| --- | --- | --- | --- |
| Univariate logistic regression analysis |  |  |  |
| Age | 1.041 | 1.006-1.078 | 0.02 |
| Infection | 2.937 | 0.947-9.112 | 0.062 |
| Cancer | 3.002 | 1.126-8.004 | 0.028 |
| AF | 3.08 | 1.155-8.215 | 0.025 |
| BUN | 1.02 | 1.008-1.032 | 0.001 |
| Phosphate | 1.463 | 1.084-1.975 | 0.013 |
| NN50 | 1.0 | 1.000-1.001 | 0.045 |
| pNN50 | 1.020 | 1.001-1.039 | 0.038 |
| Multivariate logistic regression analysis |  |  |  |
| Model I |  |  |  |
| Age | 1.045 | 1.005-1.087 | 0.027 |
| Sex | 0.418 | 0.142-1.211 | 0.108 |
| NN50 | 1.000 | 1.000-1.001 | 0.396 |
| Model II |  |  |  |
| Age | 1.044 | 1.004-1.087 | 0.033 |
| Sex | 0.418 | 0.143-1.220 | 0.110 |
| pNN50 | 1.010 | 0.988-1.032 | 0.382 |
| Model III |  |  |  |
| Age | 1.052 | 1.002-1.105 | 0.04 |
| Sex | 0.334 | 0.106-1.060 | 0.063 |
| Infection | 3.036 | 0.870-10.595 | 0.082 |
| Cancer | 2.214 | 0.734-6.681 | 0.158 |
| AF | 1.518 | 0.414-5.561 | 0.529 |
| Phosphate | 0.879 | 0.553-1.399 | 0.587 |
| BUN | 1.019 | 0.999-1.040 | 0.059 |
| NN50 | 1.000 | 1.000-1.001 | 0.594 |
| Model IIIIV |  |  |  |
| Age | 1.048 | 0.996-1.102 | 0.068 |
| Sex | 0.329 | 0.103-1.053 | 0.061 |
| Infection | 3.322 | 0.916-12.051 | 0.068 |
| Cancer | 2.311 | 0.764-6.988 | 0.138 |
| AF | 1.418 | 0.400-5.024 | 0.588 |
| BUN | 1.019 | 0.999-1.040 | 0.064 |
| Phosphate | 0.879 | 0.551-1.400 | 0.586 |
| pNN50 | 1.012 | 0.984-1.042 | 0.409 |

AF, atrial fibrillation; BUN, blood urea nitrogen; NN50, the mean number of times in which the change in successive normal sinus (NN) intervals exceeds 50 ms; pNN50, % of successive RR intervals differing >50 ms; CI, confidence interval.

However, multivariate Cox regression analysis showed that age was the only independent predictor of 30-day all-cause mortality, while the other variables including NN50 were not independent predictors of 30-day all-cause mortality.

### ROC Curve Analysis and Kaplan-Meier Curve

Figure 1 and Table 4 show the ROC curve, AUC, best cut-off value, sensitivity and specificity for the five continuous variables that significantly differed between survivors and non-survivors at 30 days after admission. The AUC for NN50 and pNN50 were 0.67 (95%CI: 0.51-0.82) and 0.67 (95%CI: 0.51-0.83), respectively. All patients were then stratified into high- and low- level groups according to the best cut-off value and Kaplan-Meier survival analysis curves were plotted for age, BUN, NN50 and pNN50 (Figure 2), which showed that the survival curves of patients in the two groups were significantly different (*P* < 0.001).

**Figure 1.**
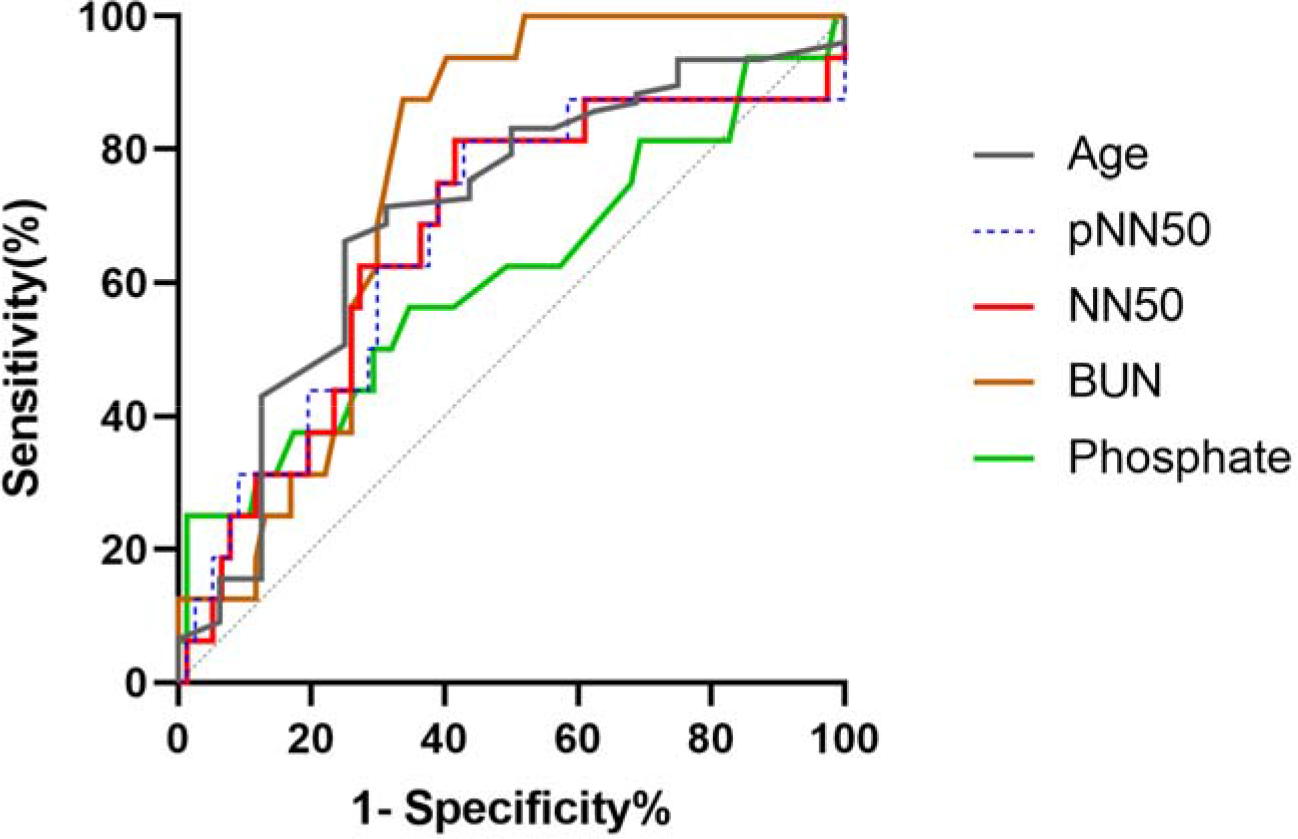
ROC curves of variables for predicting 30-day all-cause mortality. The ROC curves for the age, pNN50, NN50, BUN and phosphate are shown as gray, blue, red, orange and green lines, respectively. NN50, the mean number of times in which the change in consecutive normal sinus (NN) intervals exceeds 50 ms. pNN50, % of consecutive RR intervals differing >50 ms. BUN, blood urea nitrogen.

**Figure 2.**
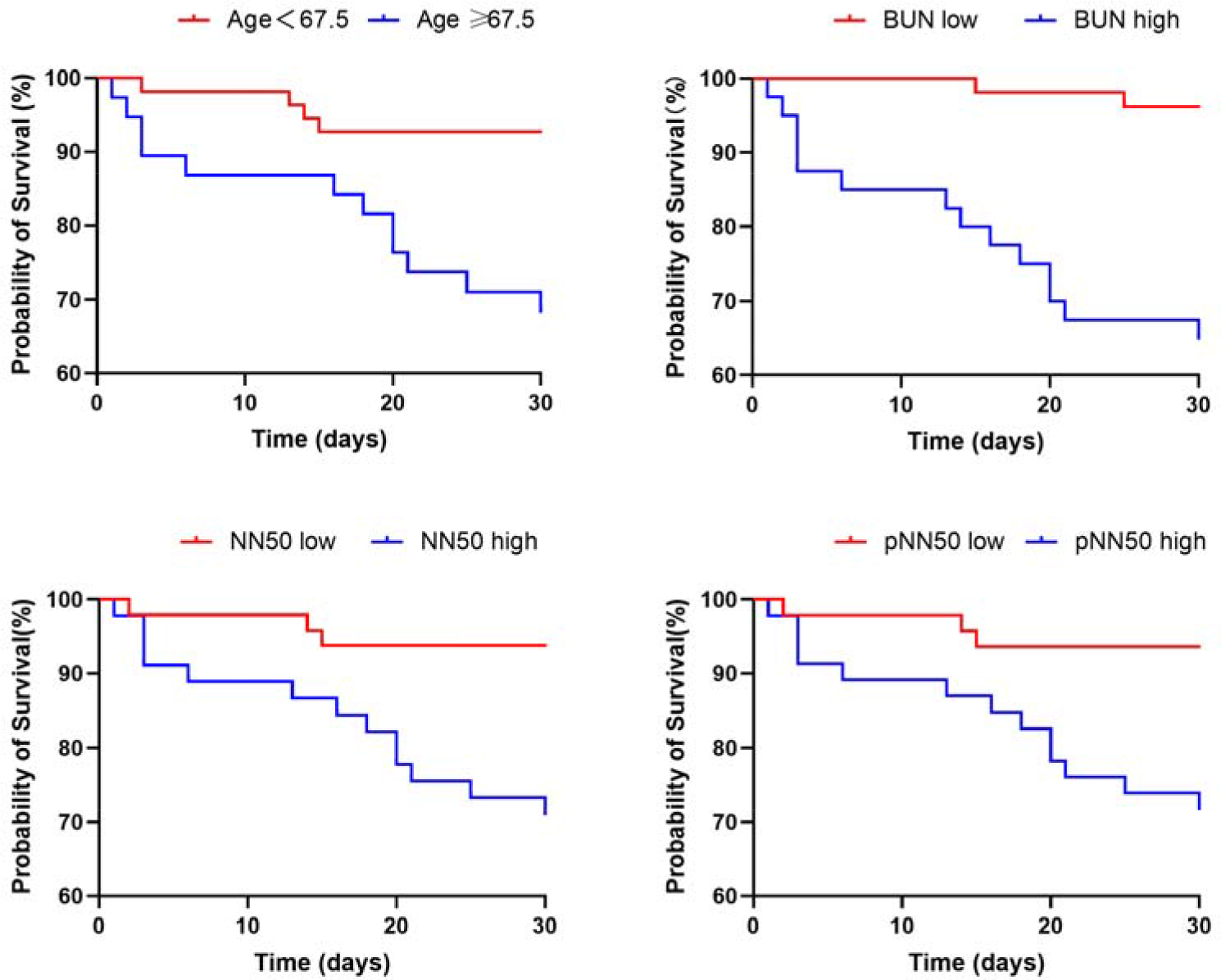
Kaplan-Meier survival curves for all-cause mortality at 30 days after admission. Log-rank test showed *P*=0.003, *P*<0.0001, *P*=0.005, and *P*=0.006 for age, BUN, NN50 and pNN50, respectively. NN50, the mean number of times in which the change in consecutive normal sinus (NN) intervals exceeds 50 ms. pNN50, % of consecutive RR intervals differing > 50 ms. BUN, blood urea nitrogen.

**Table 4.**
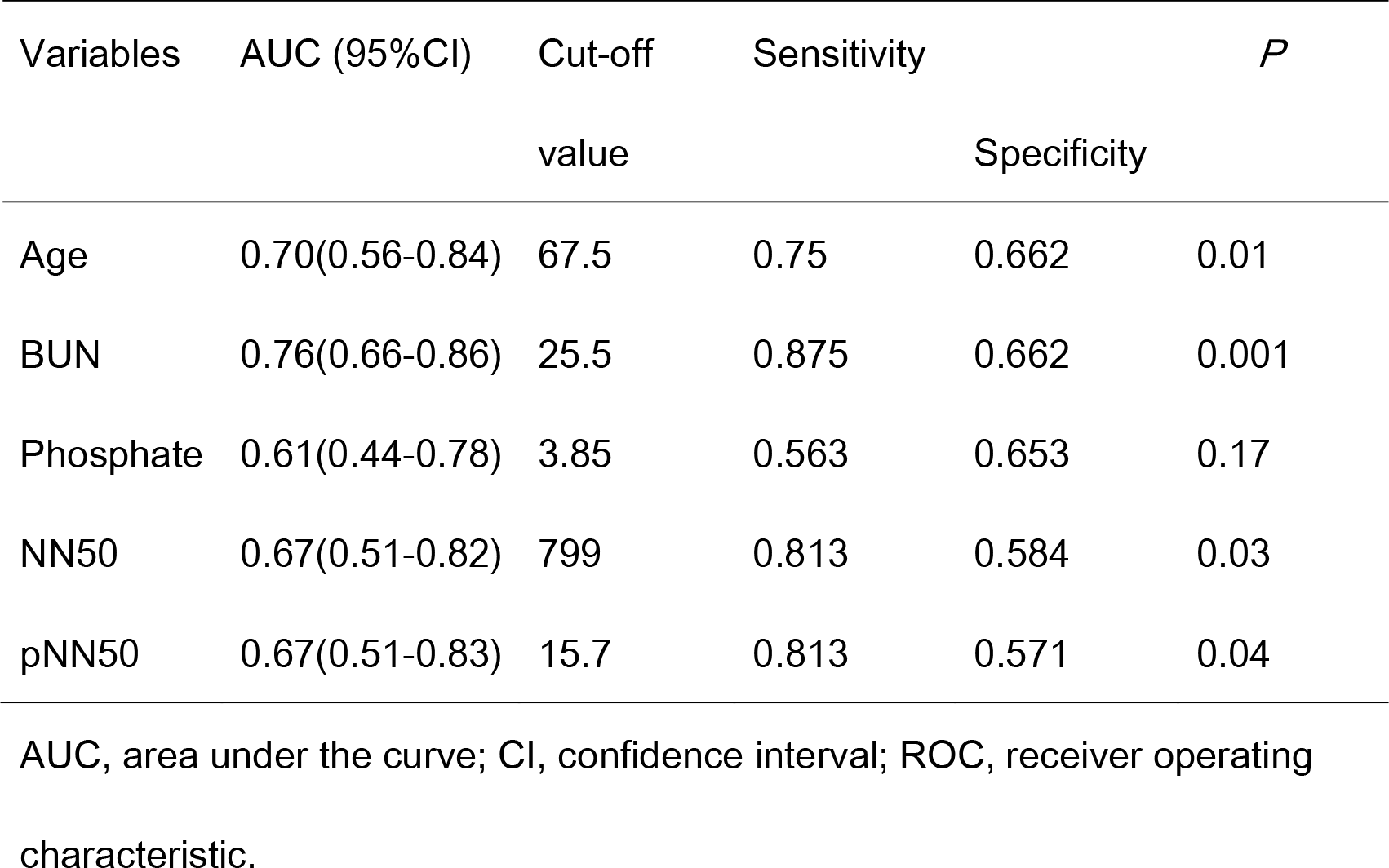
ROC curve parameters in Figure 1

| Variables | AUC (95%CI) | Cut-off<br>value | Sensitivity | Specificity | *P* |
| --- | --- | --- | --- | --- | --- |
| Age | 0.70(0.56-0.84) | 67.5 | 0.75 | 0.662 | 0.01 |
| BUN | 0.76(0.66-0.86) | 25.5 | 0.875 | 0.662 | 0.001 |
| Phosphate | 0.61(0.44-0.78) | 3.85 | 0.563 | 0.653 | 0.17 |
| NN50 | 0.67(0.51-0.82) | 799 | 0.813 | 0.584 | 0.03 |
| pNN50 | 0.67(0.51-0.83) | 15.7 | 0.813 | 0.571 | 0.04 |

AUC, area under the curve; CI, confidence interval; ROC, receiver operating characteristic.

## Discussion

The present study found that unadjusted age, presence of malignancy and AF, BUN, phosphate, NN50 and pNN50 were all associated with elevated 30-day all-cause mortality in ICU patients. However, after adjustment for covariates, NN50 or pNN50 were not independent predictors of 30-day all-cause mortality in ICU patients.

Heart rate variability (HRV) is related to the balance between the sympathetic ANS, which innervates the entire myocardium, and the parasympathetic ANS, which innervates the sinus node, atrial myocardium and atrioventricular node in the control of the heart [19]. It reflects the ability of the heart to react to numerous physiological and environmental stimuli, such as respiration, exercise, hemodynamic and metabolic changes and disease-related stress[19]. The time-domain HRV variables NN50, pNN50 and rMSSD are thought to be closely related to parasympathetic activity [20]. There is no established threshold value for these variables in healthy subjects, but previous studies have reported a range of 433-488 for NN50, 5% to 18% for pNN50 and 21-43ms for rMSSD based on 24-hour ECG recordings in healthy adult subjects[21–24]. In our study, the level of NN50, pNN50 or rMSSD in survivors was comparable to previously reported values and significantly lower than in non-survivors, suggesting an overstimulated parasympathetic tone in the non-survivors. The underlying causes for the significantly higher parasympathetic-cardiac tone in the non-survivors were unclear and could be explained by several hypotheses. First, the difference in sedative use between the two groups may explain this as sedatives are thought to affect cardiac autonomic function[25]. However, a post-hoc analysis showed no significant difference in the use of sedatives (propofol and midazolam) between both groups. Second, since baroreflex sensitivity partly reflects parasympathetic innervation [26], the higher parasympathetic activity seen in our study may be explained by higher baroreflex activation as a result of a relatively lower systolic and mean arterial blood pressure in the non-survivor group. It should be noted that direct assessment of parasympathetic and baroreflex activity is difficult to obtain in humans and there is no alternative way to validate our hypothesis within the scope of the current study. Third, previous human studies have suggested that HRV variables reflecting parasympathetic activity are inversely related to inflammatory indicators such as Interleukin-6 and C-reactive protein[27], whereas in experimental studies vagus nerve stimulation alleviates the production of proinflammatory cytokines and suppresses the inflammatory process[26]. Therefore, we speculate that the higher parasympathetic activity in the non-survivor group may be an enhanced adaptation of the parasympathetic or vagal tone as a result of the high prevalence of infection to attenuate the inflammatory process. However, due to the lack of measurements of inflammatory markers in the study subjects, we cannot be certain that inflammatory markers were significantly different between both groups and future studies are required to prove our hypothesis. The clinical implications of increased parasympathetic tone in intensive care medicine have been less reported in the literature. A study of patients with COVID-19 infection and mechanical ventilation reported a greater parasympathetic modulation of heart rate, as evidenced by higher levels of pNN50 and rMSSD compared with patients without COVID-19 infection, and the author concluded that the monitoring of parasympathetic tone in COVID-19 infection may be a predictive marker of disease progression [28]. In another study of patients with severe head injury, increased parasympathetic tone assessed by HRV on the day after trauma was predictive of brain death, whereas a decreased parasympathetic tone in the awakening period was associated with worse clinical recovery, suggesting a dynamic change in autonomic nervous tone in these patients[29]. Taken together, our study and previous studies suggest that the imbalance between cardiac sympathetic and parasympathetic tone in ICU patients may negatively affect the overall prognosis. In contrast to our study, Pontet et al. reported that a lower level of rMSSD may predict the development of MODS in septic patients [7]. It should be noted that our analysis was derived from a 24-hour ECG recording after ICU admission, whereas Pontet et al. used relatively shorter ECG recordings (10 minutes) in their study, and it is inappropriate to directly compare HRV results from different recordings.

Frequency domain analysis provides information on the contribution of different frequency components of HRV to the heart rate modulation. In general, it is proposed that SNS modulates LF oscillations, PNS affects both LF and HF oscillations, hormonal factors modulate VLF oscillations of HR and circadian rhythm affects the ULF band [3, 30]. Our study showed that non-survivors had a significantly lower level of ULF power compared to survivors at 7 days after admission, while no significant difference was noted for the other frequency components. The physiological significance of ULF power is less clear compared to VLF, LF and HF power, but may be influenced by factors such as circadian rhythm, core body temperature, metabolism and the renin-angiotensin system[31]. In the literature, several frequency-domain HRV variables have been reported to predict mortality or disease deterioration in critically ill patients. For example, reduction in VLF was predictive of mortality in patients with multiple organ dysfunction syndrome [6] whereas normalized HF power was reported to be a valuable predictor of in-hospital mortality in patients with sepsis [8]. However, Barnaby et al. found that the LF/HF ratio was not a reliable clinical predictor of in 72-hour deterioration in patients with sepsis presenting to the emergency department [32]. These studies suggest that the ability of HRV to predict prognosis may vary in patients with the same diagnosis.

Nonlinear measures of HRV provide information about the heart rate variance by analyzing temporal similarities in the signals and are suitable for non-stationary data analysis[3]. Detrended fluctuation analysis provides short-term(a1) and long-term(a2) fluctuations between R-R intervals. Our study showed that at 7 days after admission, non-survivors had a significantly lower level of DFA(a1), suggesting a reduced ability of short-term heart rate fluctuation. This is consistent with previous studies [17, 33, 34] in which reduced DFA(a1) was a strong prognostic predictor of poor clinical outcome or mortality in patients with out-of-hospital sudden cardiac arrest[17], myocardial infarction[33], or end-stage renal disease receiving peritoneal dialysis[34].

The identification of age as the only independent predictor of mortality in our study is not surprising. However, we identified a cut-off age of 67.5 years with a sensitivity of 0.75 and specificity of 0.662 to predict mortality in ICU patients. This is of clinical relevance as it is still debated whether older patients will benefit from ICU admission[35]. In a French cohort study including 133 966 patients admitted to ICUs, the risk of 3-year all-cause mortality increased progressively with age, with a sharp increase after 80 years of age[36].The relatively younger age value in our study may be explained by the relatively small sample size, as only 93 patients were recruited for the analysis out of 50920 ICU patients in the MIMIC-IV database.

Our study showed that blood urea nitrogen was the only variable that remained significantly higher in non-survivors than in survivors at all study points. More importantly, the identified cut-off value of 25.5 mg/dL (equivalent to 9.1mmol/L) for predicting mortality is comparable to cut-off values used in other risk stratification tools incorporating BUN such as the CURB criteria and the Pneumonia Severity Index for pneumonia severity[37]. Our study is in agreement with previous literature [38], in which BUN was associated with in-hospital mortality in critical patients and provides further evidence for its use in risk stratification in critical patients.

Several limitations of our study must be addressed. First, this is a single-center retrospective study with a small sample size and it should be noted that only one Asian patient was included in the study. Therefore, our findings may not be generalizable to all ICU patients, especially in Asian countries. Future studies focusing on Asian ethnicity are needed to validate our findings. Second, due to the nature of the database[10], a patient may have up to 39 diagnoses with imperfect ranking of importance for each hospitalization. Therefore, the most important reason requiring ICU stay for patients remained unclear and all patients were grouped together for the final analysis. This can be overcome in future prospective studies if the primary diagnoses of patients are accurately determined and recorded. Third, some important clinical characteristics such as height and laboratory indicators such as albumin and alanine aminotransferase were not available for all patients and were excluded from the study. Therefore, the result may be affected by these missing data. Finally, the quality of some patients’ cardiac monitoring data was affected by interferences such as body movement. Our team developed algorithms to remove these interferences as much as possible but may not be able to completely eliminate their possible influence on the final result. In addition, ideally, 24-hour cardiac monitoring starting at the same time point (e.g., 8 a.m.) should be used for all patients. However, due to the differences in admission time, analysis using the data as previously proposed would lead to a loss of important information and was therefore not considered.

## Conclusions

Our study shows that NN50 and pNN50 are associated with increased 30-day all-cause mortality in ICU patients but are not an independent predictors of all-cause mortality using multivariate COX regression analysis. Nevertheless, HRV may still be a valuable tool in predicting disease progression or mortality in ICU patients and early detection and intervention of HRV abnormalities may improve the prognosis of ICU patients.

## Supplementary Information

### Acknowledgements

The authors acknowledge the technical advice and assistance provided by Dr Junjun Fan from Shenzhen University.

### Funding source

Funding for this project were provided by the Guangdong Basic and Applied Basic Research Foundations (No. 2021A1515010402 and 2021A1515011433).

### Authors’ contributions

S.L., Z.Y. and L.J. conceived the research hypothesis, S.L., P.W., Q.Y., Z.Y. and L.J. contributed to the design of the research protocols; S.L., Q.Y., P.W., and Y.L. performed data extraction and analysis. S.L., Z.Y. and L.J. analyzed and interpreted the results of the study. S.L., Z.Y. and L.J. drafted the manuscript and prepared the figures. All authors reviewed and approved the final version of the manuscript.

### Availability of Data and Materials

All data and materials are available upon request.

### Ethics approval and informed consent

Studies involving human participants were reviewed and approved by the Institutional Review Board of the Massachusetts Institute of Technology and the Beth Israel Deaconess Medical Center. Written informed consent for participation was not required for this study in accordance with the national laws and the institutional requirements. The study was also approved by the Ethics Committee of Sun Yat-sen Memorial Hospital, Sun Yat-sen University (approval ID: SYSKY-2023-089-01).

### Consent for publication

All authors give their consent for the publication of this manuscript.

### Supplemental Material

Tables S1–S7 Figure S1

## Data Availability

All data and materials are available upon request.

https://physionet.org/content/mimiciv/2.2/

https://physionet.org/content/mimic4wdb/0.1.0/

